# Contact tracing during Phase I of the COVID-19 pandemic in the Province of Trento, Italy: key findings and recommendations

**DOI:** 10.1101/2020.07.16.20127357

**Authors:** Pirous Fateh-Moghadam, Laura Battisti, Silvia Molinaro, Steno Fontanari, Gabriele Dallago, Nancy Binkin, Mariagrazia Zuccali

## Abstract

**Introduction:** Contact tracing is a key pillar of COVID-19 control. In response to the COVID-19 epidemic in the Autonomous Province of Trento (Italy) a software was developed to standardize data collection and facilitate surveillance of contacts and outbreaks and map the links between bases and contacts. In this paper, we present the results of contact tracing efforts during Phase I of the epidemic (March-April, 2020, mostly under lockdown), including sociodemographic characteristics of contacts who became cases and of the cases who infected one or more contact.

**Methods:** A contact tracing website was developed that included components for geolocation and linking of cases and contacts using open source software. Information on community-based confirmed and probable cases and their contacts was centralized on the website. Information on cases came directly from the central case database, information on contacts was collected by telephone interviews following a standard questionnaire. Contacts were followed via telephone, emails, or an app.

**Results:** The 2,812 laboratory-diagnosed community cases of COVID-19 had 6,690 community contacts, of whom 890 (13.3%) developed symptoms. Risk of developing symptomatic disease increased with age and was higher in workplace contacts than cohabitants or non-cohabiting family or friends. The greatest risk of transmission to contacts was found for the 14 cases <15 years of age (22.4%); 8 of the 14, who ranged in age from <1 to 11 years) infected 11 of 49 contacts. Overall, 606 outbreaks were identified, 74% of which consisted of only two cases.

**Discussion:** The open-source software program permitted the centralized tracking of contacts and rapid identification of links between cases. Workplace contacts were at higher risk of developing symptoms. Although childhood contacts were less likely to become cases, children were more likely to infect household members, perhaps because of the difficulty of successfully isolating children in household settings.

## Introduction

The autonomous province of Trento in northern Italy, which has a population of 541,000, was heavily affected by the COVID-19 epidemic in Italy during March and April, 2020 (Phase 1). Despite a complete lockdown that began on March 10 with closure of schools and universities and all businesses except grocery stores, pharmacies, and newsstands, the number of cases rose exponentially through the end of March and reached a plateau in April.

Trento implemented contact tracing for cases of COVID-19 shortly after the first cases were identified in the province among three tourists from the Lombardy region (end of February). Efforts were led by the provincial agency for health services (APSS), which oversees the four local district health units and 7 provincial hospitals and provides preventive and laboratory services, and by the APSS department of prevention, which is responsible for surveillance.

Nasal swabs for suspect cases in the community were performed in various settings (clinics, drive thrus, home visits) and were tested in three provincial laboratories. Positive results were notified to the provincial department of prevention from where the case information was forwarded to the local district of residency of the case, where public health visitors, nurses and physicians then conduct detailed investigations of each case. General practitioners notified clinically suspect cases directly to the department of prevention.

Follow up through the quarantine period was provided by the contact tracers in each local health district unless 1) the contacts themselves became cases or 2) if they cohabitants of a known case. The follow up was done via phone, an app, or email based on the contact’s preference. Domestic care units provided the follow up for these two groups and were responsible for reporting any cases that developed among cohabitants to the central care database of their respective health units. Contact tracing was performed by public health visitors, nurses and doctors and, after case numbers overwhelmed capacities by other health workers such as safety inspectors, controllers or administrative personnel, who were repurposed as contact-tracers.

At the beginning of the pandemic, the contact tracing activities of the individual services were recorded non-standardized Excel sheets, which were inconsistently completed by those performing the tracing. This method of collecting information did not always provide overall information on the number of contacts under surveillance, on the nature of the relationship between case and contact, on how many contacts had in turn become cases (outbreaks), and on the starting and ending dates of follow-up surveillance. We therefore developed a computer-based monitoring system that standardized data collection and facilitates the surveillance of contacts and outbreaks, allowed periodic analyses for the production of standard reports, and permitted more detailed epidemiological analysis for better identification of high-risk contacts and the targeting of contact tracing efforts. It also allowed for the identification of any cases that developed among contacts who were cohabiting with a known case.

Although contact tracing is an important pillar of the Test, Treat and Track strategy of the World Health Organization (^1^), little is known about the yield of such tracing. In this paper, we present the results of contact tracing in the Province of Trento for March and April 2020, including number of contacts per case, secondary attack rates overall and by the demographic characteristics of the contacts, and the association between case characteristics and the likelihood of their contacts themselves becoming cases.

## Materials and methods

To standardize data collection and to build a system capable of creating a database for epidemiological analysis and accessible from the public health point of view, a contact tracing website, based on the Django framework, with extensions in Python(^2^), was developed. The system for demonstrating links between cases and contacts was developed using the visJS library (^3^). The Geodatabase component was created in PostgreSQL with PostGIS spatial extension and Georeferencing was performed using Open Street Map APIs (^4^).

The surveillance platform, which was named “COVID 19”, collected information on the contacts of confirmed and probable cases subjected to self-isolation by the local health authorities. Information on cases came directly from the central local health unit database, information on contacts was collected by telephone interviews following a standard questionnaire. The information on each person’s contacts included:

- personal data: name, surname, date of birth, gender, *unique fiscal identifier*, residence, telephone number, and email address as well as the name, surname and telephone number of their general practitioner or pediatrician, and whether the contact is a member of the general population, is in an institutionalized setting such as a nursing home, or is a health care worker.
- nature of the contact: date of the last contact with the case to which the contact is connected and type of relationship between contact and case (cohabitant, family member or friend not living with the case, work colleague, other)
- Follow up and monitoring: preferred method of ongoing contact (telephone, email, app), start and end date of monitoring, onset of symptoms (fever greater than 37.5 degrees and / or persistent dry cough) and date of onset of symptoms, and transfer to domestic care services for those with symptoms and date of transfer. For the contacts who were cohabitants of cases and who were transferred for follow up to domestic care services, information on whether they had become cases themselves was ascertained by review of the case-reporting database.

According to ECDC and governmental guidelines^5^ a contact of a COVID-19 case has been considered any person who has had contact with a COVID-19 case within a time frame ranging from 48 hours before the onset of symptoms of the case to 14 days after the onset of symptoms.

On the platform, each contact was linked to its reference case. The information collected on the case included whether it was a laboratory-confirmed or probable case based on symptoms following contact with a known case. It was decided not to collect additional information on the cases in anticipation of future linkage with the case database, which was managed separately.

In addition to data collection, the platform allowed the download of data to Excel and mapping of the relationships between the cases and their contacts, as show in Figure 1. Such immediate visualization was useful for the identification of possible outbreaks to be investigated.

**Figure 1.**
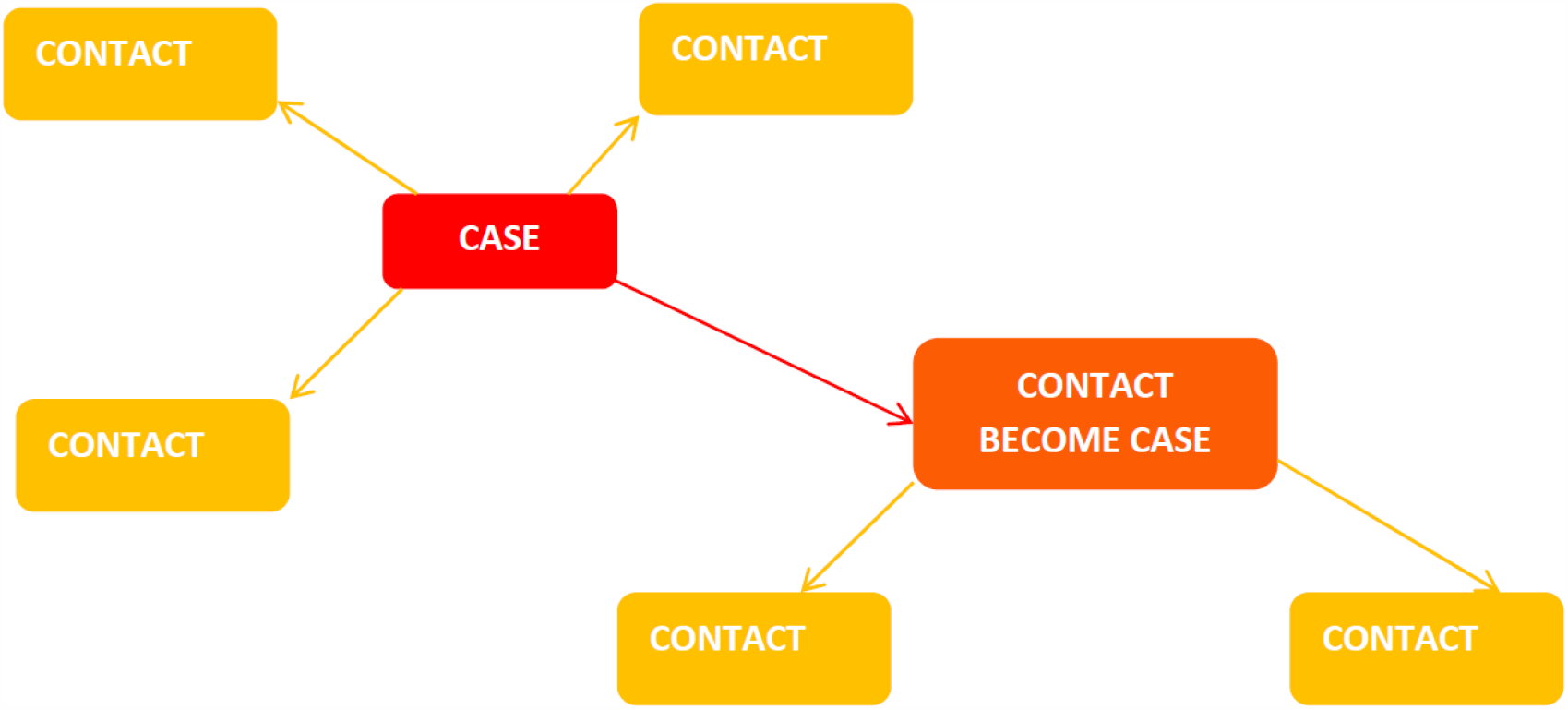
Map of relationship between contacts.

We used the SAS Enterprise Guide 7.12 to analyze the data from the COVID-19 contact tracing platform for the months of March and April 2020. We examined follow up outcomes and the secondary attack rates overall and by the demographic characteristics of the contacts. We also examined the association between case characteristics and the likelihood of their contacts themselves becoming cases. For purposes of the analysis, contagiousness was defined as the percentage of the contacts of a case becoming cases themselves. Finally, we examined the number and size of outbreaks, using the definition that 2 or more related cases constitute an outbreak.

## Results

During March and April 2020, 7,791 persons were identified as contacts and placed in self-isolation. Of these, 1,101 were linked with institutional settings, including nursing homes (898 contacts in 31 structures), hospitals (158 contacts), day and residential centers for the disabled and similar structures (26), and convents (19). These contacts have been excluded from the current analysis.

Of the remaining 6,690 contacts, 6,577 (98.3%) were contacts of cases residing in the province of Trento and 113 (1.7%) were contacts of cases who lived elsewhere.

The contacts were linked to 2,812 cases (mean 2.3 contacts per case; median 1; range 1 to 42). Thirty percent of the cases had no identified contacts, and nearly half had between 1 and 3 contacts. Of the 2,812 cases, 1,979 (70.4%) cases were laboratory confirmed.

### Characteristics of contacts

Contacts ranged in age from 0 to 110 years (median 43). Approximately half (50.7%) were men. Overall, a total of 56.0% were living in the same household as cases, 27.2% were non-cohabiting family or friends, 8.0% were workplace contacts and 8.8% were other. However, these proportions differed over time as shown in Figure 2. Data were not consistently available on type of contact during the first week of March. During the first week for which data were available (March 8-14), non-cohabiting family and friends constituted the largest group of contacts. After the national lockdown took place on March 10, cohabitants became the predominant type of contact, accounting for 2/3 of the contacts. The mean number of contacts traced per case also diminished after the lockdown, from 4.1 contacts per case (median: 2; range 0-37) during the week of March 8 to a low of 2.1 (median: 1; range 0-7) during the week of April 26.

**Figure 2.**
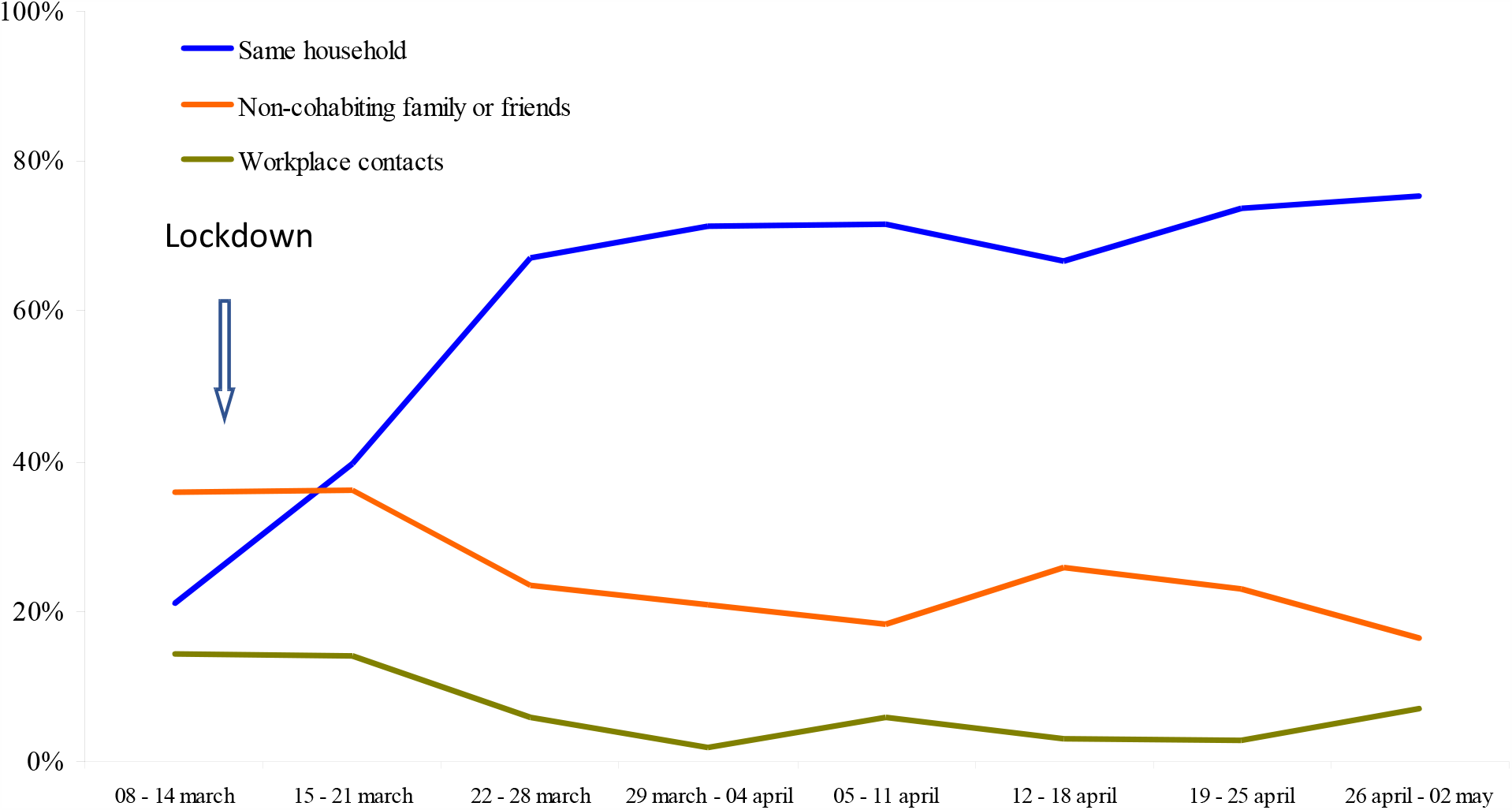
Distribution of type of contact by week, Province of Trento, March-April 2020 (n=5,252)

### Follow up information

A total of 3,351 (56.2%) of the 6,690 contacts were transferred over to the home care monitoring team because they developed symptoms (890) or because they were cohabiting with a case (2461). The remaining 2,999 (44.8%) completed public health surveillance without developing symptoms. Data on end date of surveillance or transition to the home care monitoring team was not available for 340 contacts (5.1%).

### Secondary attack rates

Of the 6,690 contacts, 890 developed symptoms after contact with a confirmed case and themselves became cases, yielding a secondary attack rate of 13.3%. A total of 485 (54.5%%) of the 890 contacts who became cases were laboratory-confirmed; the remaining 405 (45.5%) were defined as probable cases based on symptoms and their epidemiologic link with a case. The number of contacts, the number who became cases, and the secondary attack rate by age, gender, and type of contact are shown in Table 1. The risk of developing symptoms or being found to have a positive test and thus being defined as a case increased with the age of the contact, from a low of 8.4% in contacts 0-14 years of age to 18.9% in those over 75 years. There was no major difference by gender. Workplace exposure was associated with higher risk of becoming a case than cohabiting with a case or having a non-cohabiting family member or friend who was a case.

**Table 1.** Percentage of contacts who were became cases, by age, gender, and type of contact. Public Hygiene Services, province of Trento - March-April 2020

| Characteristic of contact | #of contacts | # of contacts who became cases | Secondary AR |
| --- | --- | --- | --- |
| **Age, years (n=6,687)** |  |  |  |
| 0-14 | 1,024 | 86 | 8.4% |
| 25-29 | 1,372 | 126 | 9.2% |
| 30-49 | 1,646 | 245 | 14.9% |
| 50-64 | 1,712 | 264 | 15.4% |
| 65-74 | 467 | 79 | 16.9% |
| 75+ | 466 | 88 | 18.9% |
| **Gender (n= 6,406)** |  |  |  |
| Women | 3,156 | 426 | 13.5% |
| Men | 3,250 | 427 | 13.1% |
| **Nature of contact with case (n=6,255)** |  |  |  |
| Cohabitant | 3,546 | 500 | 14.1% |
| Non-cohabiting family or friend | 1,596 | 206 | 12.9% |
| Work colleague | 499 | 79 | 15.8% |
| Other | 614 | 55 | 9.0% |

**Table 2.** Contagiousness of index cases by age and gender, Province of Trento - March-April 2020.

| Characteristic of case | Cases | #of contacts | # of contacts who became cases | Contagiousness |
| --- | --- | --- | --- | --- |
| **Age, years (n=1,489)** |  |  |  |  |
| 0-14 | 14 | 49 | 11 | 22.4% |
| 25-29 | 118 | 475 | 62 | 13.1% |
| 30-49 | 446 | 2,361 | 250 | 10.6% |
| 50-64 | 477 | 2,222 | 303 | 13.6% |
| 65-74 | 181 | 559 | 85 | 15.2% |
| 75+ | 253 | 909 | 155 | 17.1% |
| **Gender (n= 1,442)** |  |  |  |  |
| Women | 727 | 3,427 | 414 | 12.1% |
| Men | 715 | 2,973 | 416 | 14.0% |

### Risk of contacts becoming cases (contagiousness) by index case characteristics

Of the 2,812 cases, 433 (15.4%) reported zero contacts and were eliminated from the analysis, Additionally, the 890 contacts who became cases were not considered as cases but were instead included among the contacts for the calculation of contagiousness. Contagiousness was thus examined for 1,489 total cases.

The age of the cases for whom contagiousness could be calculated ranged from 0 to 110 years, with a median of 55 years. The number of males and females were virtually identical. Among the 14 cases 0-14 years of age, 11 of their 49 contacts became cases (22.4%), the highest rate of contagiousness of any age group (N.B. schools were closed during the two study months). In this age group, 8 of the 11 cases infected others; of those who were associated with secondary cases, three were <5 years, four were between 5 and 10 years, and one was 11 years of age. The lowest rate of contagiousness was found among cases who were 30-49 years of age (10.6%), with rates in those over 50 years increasing with higher ages. The rates of contagiousness were somewhat higher for men than for women (14.0% versus 12.1%).

### Outbreaks

During the months of March and April 2020, 606 outbreaks occurred in the province of Trento. The vast majority of outbreaks were made up of 2 connected cases (73.8%), with 16.3% having 3 cases, 6.8% having 4 and the remaining 3.1% having 5 or more connected cases (Maximum of 8 cases; figure 3).

**Figure 3.**
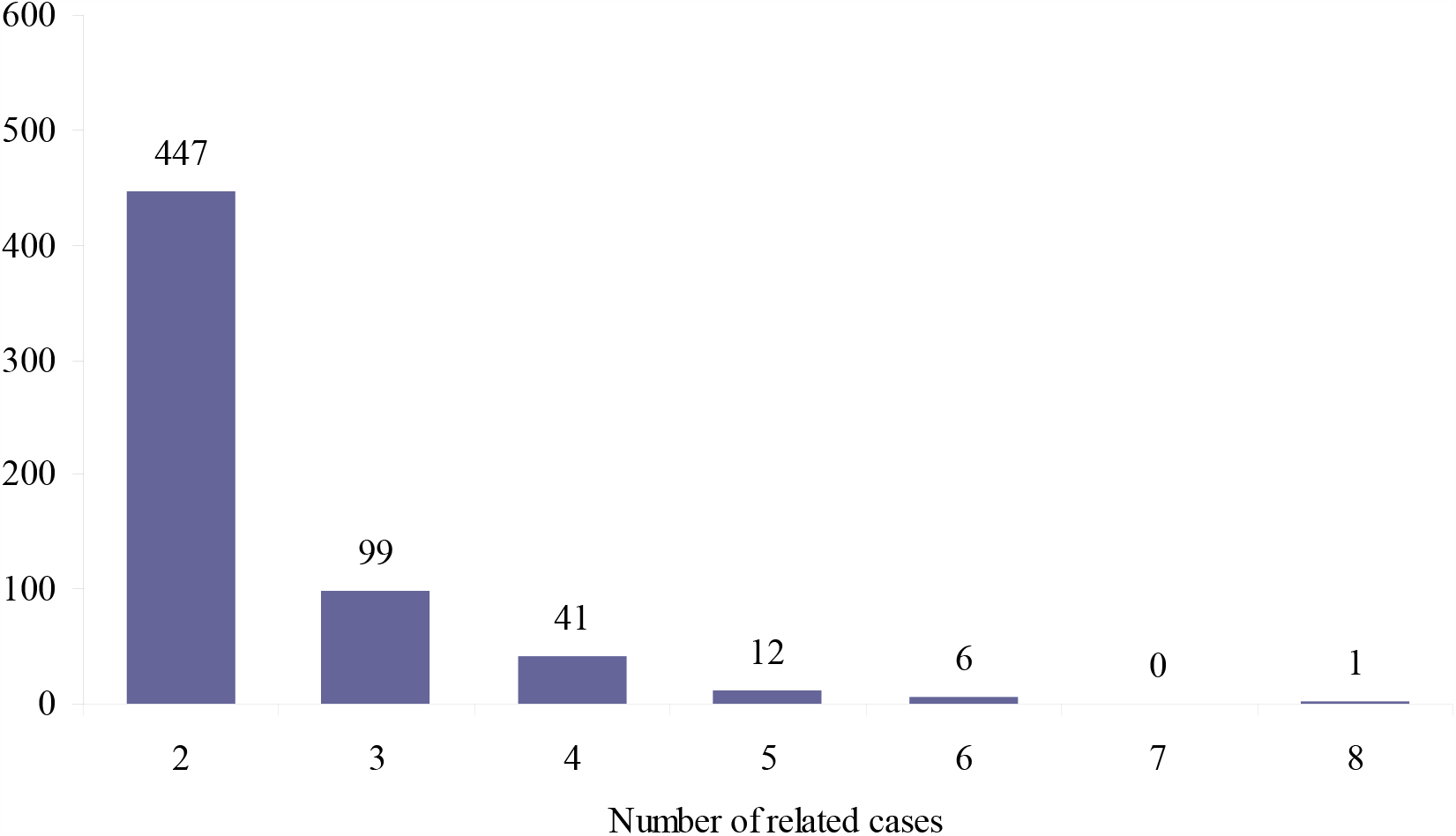
Number of outbreaks by size.

### Interval between date of last contact between case and contact and first day of active contact follow up

Dates of last contact with the case and initiation of follow up were available for 3,826 of the 6,690 contacts (57.2%). Of the remainder, 2,012 were missing the date of the last contact with the case, 349 the start date of surveillance, and 503 were missing both. The majority (69.5%) of those missing the date of last contact were cohabitant contacts whose follow up had been transferred to social services.

On average, 3.8 days (0 to 48 days; median 2 days) elapsed from the last contact to initiation of follow up. However, as shown in Figure 4, the total number of contacts to be traced increased, the interval between contact and initiation of follow-up also increased, reaching a maximum of 5.3 days during the week of March 15-21, when cases in Trento reached their peak.

**Figure 4.**
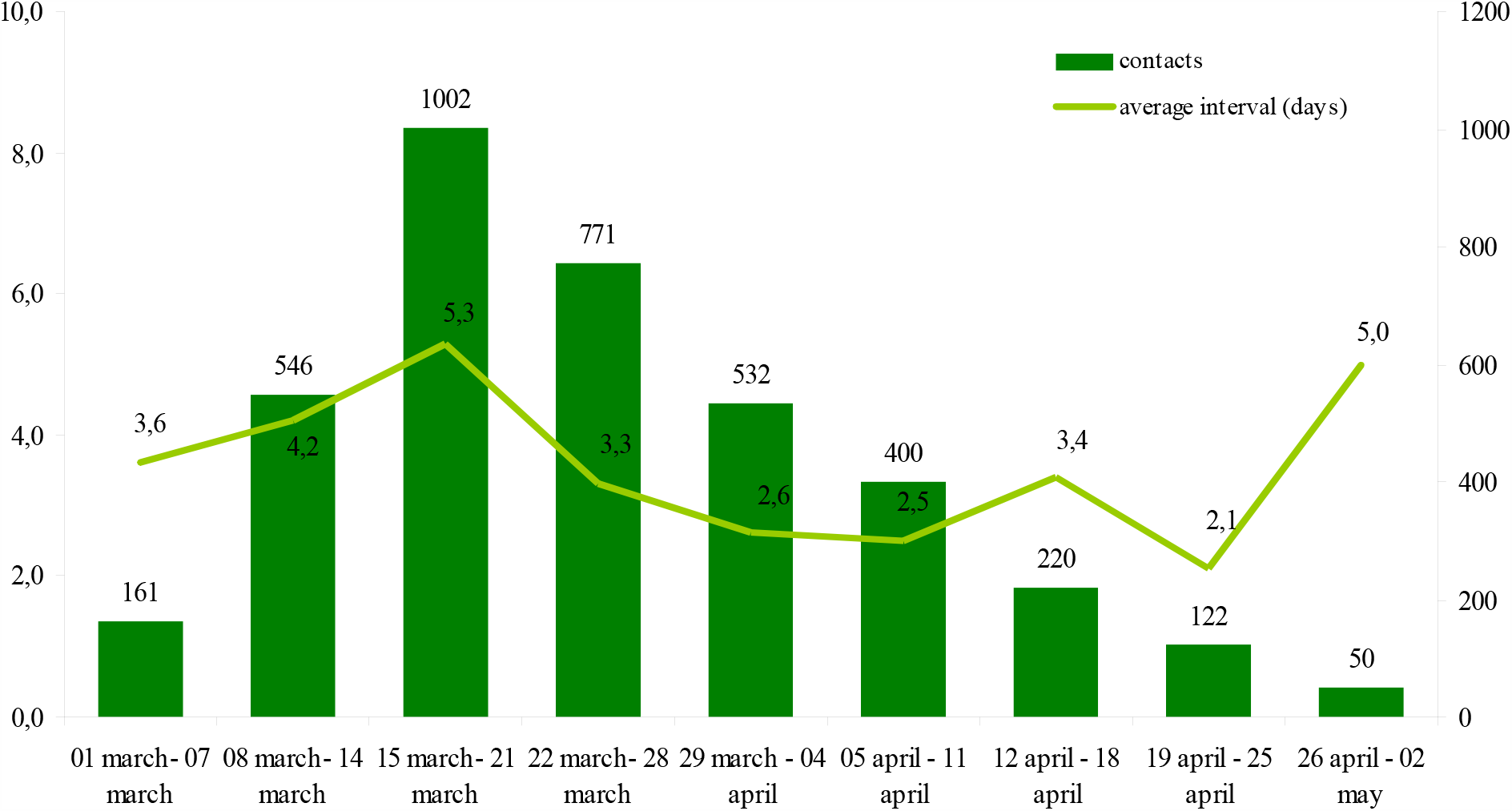
Number of weekly contacts and average days elapsed between last contact between case and contact and initiation of contact follow up, Province of Trento, March-April 2020 (n =3,826)

## Discussion

The analysis of data on contact tracing conducted by the public health services of the Province of Trento for community-diagnosed COVID-19 cases was made possible through the use of a unique locally-created surveillance platform “Covid19”. It allowed calculation of the secondary attack rate overall and by age, gender and the relationship between cases and contact, as well as providing useful information on trends and timeliness of placing contacts in self-isolation, which is important to reduce further risk of contagion.

The value of 13.3% of the secondary attack rate and 14.1% in cohabitants in our setting is comparable to what was found in Shenzen, China, where, excluding as in our analysis the cases without sufficient information, the overall secondary attack rate was 9.7% and was 14.9 % among cohabitants (^6^) A first analysis of US data reported a secondary attack rate between cohabiting partners of 10.5% (^7^).

From our analysis, children who are contacts have a lower risk of developing symptoms. This result is in line with what was reported by Qifang Bi (^6^), and by Zhang et al (^8^) in China which estimates the risk of infection of children to be significantly lower than that of adults, with the highest risk seen in the elderly.

The role of transmission from children to others is an area of great interest. The analysis carried out at the Charité laboratory in Berlin on 3,700 positive swabs processed from January to April 2020 showed that the viral load of children is similar to that of adults, confirming that children can also transmit the virus (^9^).

Indeed, in our study, children 0-14 years had a higher risk (22.4%) than any other age group of passing the infection on to others. Of particular note was the young age of the children in the study who had transmitted the disease, all but one of the 8 children who had one or more contact meeting the COVID-19 case definition were less than 10 years old, and three were under the age of 5 years. This greater risk of spread resulting from contact with an infected child that emerged from our analysis might be explained by the different nature of interactions between adults and children. While the positive adult would be likely to be more adherent with isolation precautions, it may be more difficult to truly isolate children, resulting in continuing contact with parents and siblings. Overall, our data are therefore in support of a policy of maximum caution with respect to the reopening of children’s communities and primary schools.

While age affected the risk of acquiring the infection among contacts, gender did not, confirming the national data collected by the Istituto superiore di sanità (^10^) that has highlighted similar infection rates in the face of gender-diverse mortality rates. However, in our study, male Covid cases seemed to be slightly more likely to infect others than female cases for reasons that cannot be inferred from the data we collected. Greater scrupulousness in maintaining precautions such as handwashing on the part of infected women may be a consideration.

Also noteworthy in our study is the high risk of infection associated with contacts in the workplace. This was an infrequent method of contact in the lockdown period, which covered 7 of the 8 weeks of our study, but it will be of fundamental importance in the Phase 2 of reopening. While in the United States and elsewhere, workplace epidemics of COVID-19 has become a major issue (^11^,^12^),in other countries, the public debate often focuses on the most visible aspects of spacing measures, such as gatherings in parks, recreational areas of the city, etc. What happens inside factories and companies likely to be of considerable importance to controlling the epidemic.

Our study has several limitations. First, becoming infected and being identified as a case are not synonymous. Contacts were not routinely tested and in most instances, determining if they had become a case was based on symptoms plus an epidemiological link. It could be, for example, that children and young adults may be less likely to have symptoms than adults and we may have under-estimated secondary attack rates in the younger age groups. Second, in our evaluation of contagiousness, the number of cases among children was relatively small, as was the number of contacts because schools were closed during all but one week of our study interval. Nonetheless, the findings are intriguing and merit further analyses in settings where comprehensive data on cases and contacts can be adequately linked.

Although diligent efforts were made to trace contacts, it should be noted that these efforts did not succeed fully in controlling community spread and became increasingly difficult as the epidemic peaked. By April 30th, the province had cumulative case rate of 761 cases/100 000, more than twice the national rate of 341/100 000. Fortunately, the province was able to increase ICU beds by 20%, and existing structures were converted into COVID-19 hospitals, allowing the health care system to cope despite the high case rates (^13^). The ongoing spread in the face of good follow up of contacts could potentially be attributed to the importance of asymptomatic cases in disease dissemination, the lack of unified approach to hospitalized, domestic care and public health cases in a standardized contact tracing policy, issues in case investigations during the peak of the epidemic, and delays in initiating contact tracing.

These findings suggest the need for an integrated data system where case information and laboratory results can be instantly notified, case investigations begun in a more timely way, and thus contact tracing also implemented earlier in the course of illness when the contacts may be transmitting to others. Surge capacity will be needed, however, since as the number of cases increases, the burden of work is likely to become rapidly overwhelming.

In conclusion, the combination of physical spacing measures (lockdown), closure of non-essential activities and identification of cases and contacts with consequent treatment, and isolation and quarantine in Italy during Phase 1 has allowed us to flatten the epidemic curve and then to push it down to the point where the reproductive number R (t) was below the value of 1. A gradual reopening of activities and movements therefore became possible (Phase2), and in Italy began on May 4.

In Phase 2, the protection deriving from the lockdown is lacking and consequently, alongside individual adherence to hygiene and physical distancing recommendations, the only real possibility to block the chains of infection consists of selective and timely isolation of new cases and quarantine of their contacts. This urgent need of contact tracing underlines, once again, the fundamental importance of strengthening the services dedicated to this resource-intense activity to effectively counter the re-emergence of possible outbreaks in Phase 2 and to avoid an otherwise very likely second epidemic wave. This will be particularly important as the number of contacts per case increases as people have greater freedom to move about. It is therefore necessary, on the one hand, to strengthen the services with staff capable of carrying out tracing and surveillance activities, on the other, to adopt standardized protocols in order to direct the effort according to clearly defined objectives of knowledge, monitoring and action. Furthermore, contact tracing and monitoring activities must be facilitated and supported through suitable monitoring tools, which facilitate both contact management and timely epidemiological analysis.

## Data Availability

Attribution-NonCommercial-NoDerivatives 4.0 International (CC BY-NC-ND 4.0)

## Acknowledgments

The authors wish to thank all the contact tracers of the local health unit of Trento for their work and dedication to public health.

## Notes

### Competing Interest Statement

The authors have declared no competing interest.

### Funding Statement

No funding

### Author Declarations

The Ethics Committee of the Local Health Unit of the Province of Trento on the 3rd of July 2020 reviewd the ppaer and came to the conclusion, that given the nature of the study (retrospective analyses of a database in order to improve the quality of the health service) it does not fall under its competence. The minutes of the meeting are available from the authors.

